# Projections for first-wave COVID-19 deaths across the US using social-distancing measures derived from mobile phones

**DOI:** 10.1101/2020.04.16.20068163

**Authors:** Spencer Woody, Mauricio Tec, Maytal Dahan, Kelly Gaither, Michael Lachmann, Spencer J. Fox, Lauren Ancel Meyers, James Scott, The University of Texas at Austin COVID-19 Modeling Consortium

**Affiliations:** University of Texas at Austin; Santa Fe Institute; The University of Texas at Austin

## Abstract

We propose a Bayesian model for projecting first-wave COVID-19 deaths in all 50 U.S. states. Our model’s projections are based on data derived from mobile-phone GPS traces, which allows us to estimate how social-distancing behavior is “flattening the curve” in each state. In a two-week look-ahead test of out-of-sample forecasting accuracy, our model significantly outperforms the widely used model from the Institute for Health Metrics and Evaluation (IHME), achieving 42% lower prediction error: 13.2 deaths per day average error across all U.S. states, versus 22.8 deaths per day average error for the IHME model. Our model also provides an accurate, if slightly conservative, assessment of forecasting accuracy: in the same look-ahead test, 98% of data points fell within the model’s 95% credible intervals. Our model’s projections are updated daily at https://covid-19.tacc.utexas.edu/projections/.

## 1 Introduction

On March 26, 2020, the Institute for Health Metrics and Evaluation (IHME) at the University of Washington released a [website](https://covid19.healthdata.org/united-states-of-america) that forecasts coronavirus disease (COVID-19) healthcare demand and mortality for all states in the United States. After being cited during a White House briefing on COVID-19 modeling efforts, their forecasting model, described in a [preprint](https://www.medrxiv.org/content/10.1101/2020.03.27.20043752v1) on medRxiv [IHME et al., 2020], has received an enormous amount of attention from both the general population and scientific community. IHME has since updated the model several times resulting in considerable revisions to the COVID-19 forecasts.

The IHME approach departs from classic epidemiological modeling. Rather than using systems of equations to project the person-to-person transmission of the virus, the model postulates that COVID-19 deaths will rise exponentially and then decline in a pattern that roughly resembles a bell curve (i.e., normal distribution). The model assumes that the shape of the curve will be curtailed by social distancing measures. Key inputs driving this component of the IHME model include the reported dates of state-wide shelter-in-place orders and shapes of COVID-19 epidemiological curves observed in Chinese and European cities following the implementation of similar measures.

In light of the popular appeal of the IHME model and considerable scrutiny from the scientific community, we have developed an alternative curve-fitting method for forecasting COVID-19 mortality throughout the US. Our model is similar in spirit to the IHME model, but different in two important details.

1. For each US state, we use local data from mobile-phone GPS traces made available by SafeGraph to quantify the changing impact of social-distancing measures on “flattening the curve.” SafeGraph is a data company that aggregates anonymized location data from numerous applications in order to provide insights about physical places. To enhance privacy, SafeGraph excludes census block group information if fewer than five devices visited an establishment in a month from a given census block group.
2. We reformulated the approach in a generalized linear model framework to correct a statistical flaw that leads to the underestimation of uncertainty in the IHME forecasts.

The incorporation of real-time geolocation data and several key modifications yields projections that differ noticeably from the IHME model, especially regarding uncertainty when projecting COVID-19 deaths several weeks into the future.

## 2 Model overview

At a high level, our model shares some key properties of the IHME model.

### Similarity 1: a statistical curve-fitting approach

Ours is not an epidemiological model, in the sense that we do not try to model disease transmission, nor do we use or attempt to estimate underlying epidemiological parameters like the basic reproductive rate or attack rate. Rather, our model is purely statistical: we are fitting a curve and a probabilistic error model to observed death rates in a state, and we are extrapolating from that curve. The advantage of this approach is that it does not require estimates of critical epidemiological parameters, some of which remain elusive. The disadvantage is that it cannot project longer-term epidemiological dynamics beyond the initial wave of mitigated transmission. For this reason, we do not use the model to make projections beyond a moderate (2-3 week) horizon.

### Similarity 2: time-evolving Gaussian curves

The family of curves we use for expected deaths over time is identical to that of the IHME model. Specifically, we assume that expected daily death rates can be *locally* approximated by a three-parameter curve that is proportional to a Gaussian kernel. This approximation is local in the sense that the curve’s three parameters are allowed to evolve in time as a function of state-level covariates. Just as in the IHME model, this results in fitted death-rate curves that, when plotted over time, can differ substantially from the shape of a single Gaussian. While epidemic curves do not resemble Gaussian curves, *time-evolving* Gaussian curves do provide a good fit to observed COVID-19 state-level death rates.

### Similarity 3: regression on social-distancing covariates to inform projections

As in the IHME analysis, our regression model connects each state’s deathrate curve to covariates that describe social distancing within each state. Changes in each state’s social-distancing covariates can “flatten the curve” by changing the peak death rate, the timing of that peak, and the deceleration in death rate near the peak. The strength of this approach is that it can leverage readily available data on social distancing without requiring a mechanistic transmission model. However, our model differs from the IHME model in at least three key ways, which collectively result in different forecasting behavior.

### Difference 1: real-time daily social-distancing data

We use data on Americans’ actual social-distancing behavior, derived from GPS traces from tens of millions of mobile phones across the country. This data source quantifies two main types of distancing behavior: 1) changes in visitation patterns to public places like restaurants, bars, schools, parks, pharmacies, grocery stores, etc.; and 2) time spent at home versus at work. The IHME model, by contrast, uses a much coarser measure of social distancing: the timing of state-level policy implementations like school closures and stay-at-home orders. But the mobile-phone data reveals substantial differences among states in the timing and extent of peoples’ actual distancing behavior, even for states with nominally similar policies. In Texas, for example, many large cities issued their own stay-at-home orders before the state did, affecting the movement patterns of many millions of people days before a statewide policy was in place—a fact that is clearly visible in the data. Our measures capture this substantial state-level and temporal variation that is obscured by regressing only on policies.

### Difference 2: U.S. data only

The IHME model assumes that data on death rates and social distancing policies in other countries (specifically China, at least in the original formulation of the model) can inform U.S. state-level forecasts. For a variety of reasons, we find this assumption problematic. Our forecasts therefore rely solely on U.S. data, with state-level parameters shrunk toward a common mean in a hierarchical Bayesian model.

### Difference 3: valid uncertainty quantification

We address a problem with the IHME model by relying on a fundamentally different statistical assumption about model errors. Briefly: the IHME model fits *cumulative* death rates using a least-squares-like procedure on the log scale and applying standard large-sample statistical theory to get confidence intervals. For this procedure to result in valid uncertainty quantification, one must assume that successive model errors are independent. But in the IHME fitting procedure, this assumption is violated: today’s cumulative death rate is yesterday’s plus an increment, so the two must be correlated. Our model repairs this problem by fitting *daily* (noncumulative) death rates using a mixed-effects negative-binomial generalized linear model.

## 3 Model details: a negative-binomial mixed-effects GLM for daily COVID-19 deaths

### 3.1 Model structure

We let *i* index the geographic area; in our analysis this is U.S. states, but it could be at any level of spatial resolution (e.g. country, city, etc). To make our results comparable to the IHME model, let *t* denote the number of days elapsed since a threshold death rate of 3 per 10 million residents was reached in a given area. Thus *t* doesn’t represent calendar days, but rather a notion of “epidemic days.”

Let *y*_*it*_ denote observed number of deaths in area *i* at time *t*. Let 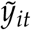 denote per-capita death rate. The IHME model assumes that the expected daily death rate *λ*_*it*_ can be locally approximated by a curve proportional to a Gaussian kernel:

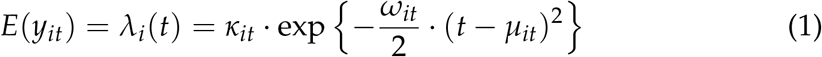

This leads to the following interpretation of the parameters:

- *κ* is the maximum daily expected death rate
- *µ* is the day on which the expected death rate achieves its maximum
- *ω* is a steepness parameter: higher *ω* means the death rate rises more rapidly as *t* approaches *µ*, and also falls more rapidly on the far side of *µ*. Specifically, the slope at the inflection point of the death-rate curve is 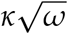.

Equation (1) expresses the most general form of the IHME model, where *θ*_*it*_ = (*κ*_*it*_, *µ*_*it*_, *ω*_*it*_) changes from area to area and day to day. This is highly overparametrized; it is necessary to enforce some type of shrinkage on these parameters in order to make the model identifiable. We address model overparametrization in a similar way to the original IHME analysis (i.e. via a hierarchical model). But we use very different data based on mobile-phone GPS traces that quantify actual distancing behavior, rather than the timing of state-level social-distancing policies (e.g. school closures, stay-at-home orders, etc.).

### The IHME model-fitting process

We note that the IHME model parameterizes its Gaussian curves in a slightly different way, but the underlying family is identical to Equation (1), in the sense that there is a bijection between our parameterization and theirs. Briefly, the IHME model assumes that the *cumulative* death rate is proportional to Gaussian CDF, and they fit the three model parameters by optimizing a penalized least-squares objective on the log-cumulative-deaths scale, interpreting the result as a Bayesian maximum a posteriori (MAP) estimate under an assumed prior. More specifically, let *N*_*i*_ be the population in state *i* and define 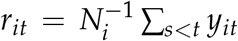 be the per-capita cumulative death rate in state *i*. The IHME model assumes that

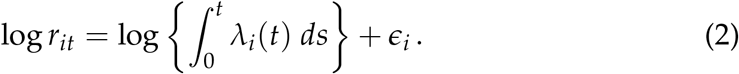

The model is fit by a penalized nonlinear least-squares procedure, encouraging the parameters of each state’s *λ*_*i*_(*t*) curve to be shrunk towards a common *conditional* mean, given social-distancing covariates. They interpret the result as a hierarchical Bayesian MAP estimate under an assumed prior, and use the inverse Hessian matrix at the MAP estimate as plug-in estimate of the posterior covariance matrix for the model parameters.

In the Appendix, we describe why this procedure does not result in valid confidence intervals. Briefly, it ignores the problems of heteroscedasticity and intra-day correlation in model errors associated with fitting to cumulative data. Together these have major consequences for uncertainty quantification. Figure 1 briefly illustrates the problems that can arise: it shows the IHME projections for Italy and Spain on April 5 looking ten days ahead, together with 95% prediction intervals. The actual data fall noticeably outside the model’s claimed range of uncertainty. This is especially worrisome, given that peak daily death rates in Italy and Spain seem to be well-characterized by the data itself through April 16th, versus the situation in many U.S. states that have yet to reach their peak, where we must rely on a model to extrapolate the location of the peak.

**Figure 1:**
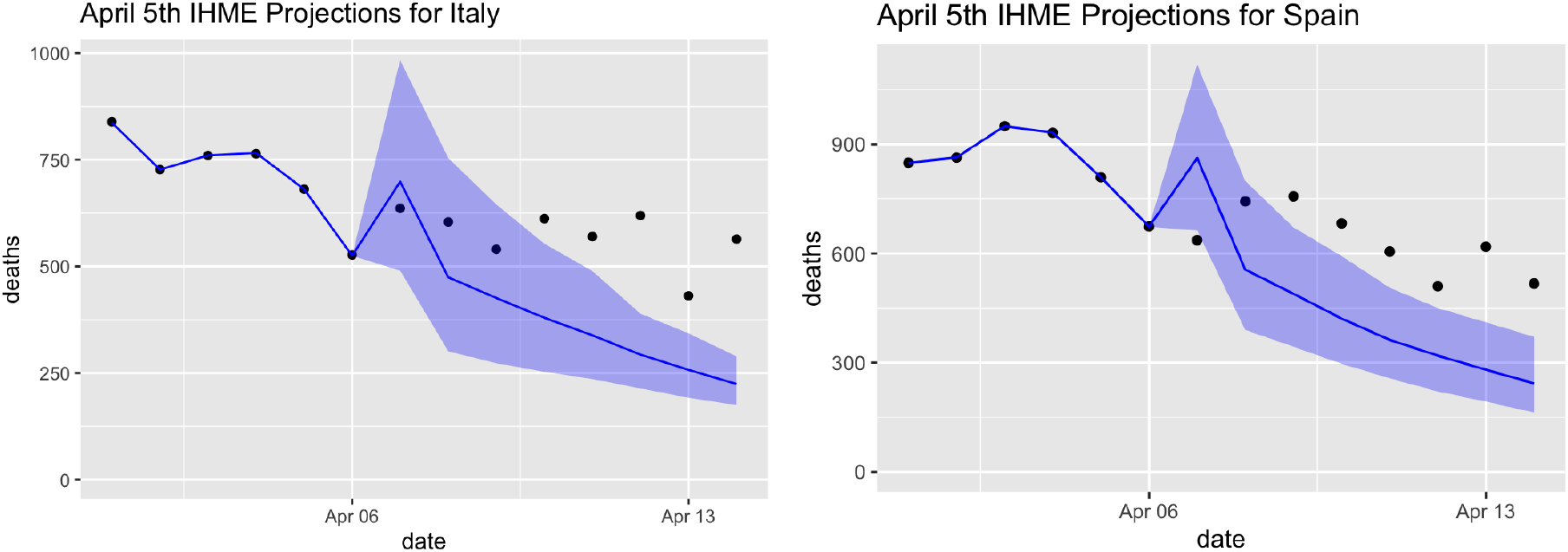
The IHME model’s April 5 projections ten days ahead for Italy and Spain (blue lines). The actual data (black dots) fall noticeably outside the model’s 95% error bars (shaded blue region), illustrating the model’s underestimate of forecasting uncertainty, even in countries whose peak daily death rates seem to be well characterized by the data.

An April 8 technical report by Marchant et. al points to similar problems with the IHME’s U.S. projections [Marchant et al., 2020]. They found that, in evaluating the model’s U.S. projections made on April 1, only 27% of the actual data points on the subsequent day actually fell within the 95% confidence bands. Luckily, this is an easily correctable problem, by placing the model in Equation (1) into the framework of generalized linear modeling. This allows for better uncertainty quantification, as we describe in the next section.

### 3.2 Our fitting approach

Observe that if we move to the log scale, Equation (1) becomes:

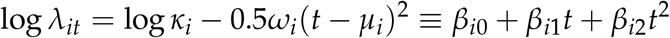

for some *β* vector. Keeping in mind that *λ*_*it*_ is the expected value for a count outcome (daily deaths *y*_*it*_), we recognize this as the expression for the linear predictor in a generalized linear model (GLM) with a log link function, as might arise in a Poisson or negative-binomial regression model for *y*. On the right-hand side, we have a locally quadratic regression on *t*, the number of elapsed days since deaths crossed the threshold value of 3 per 10 million. Moreover, there is a simple relationship between the regression coefficients *β* a nd t he original parameters of the curve:

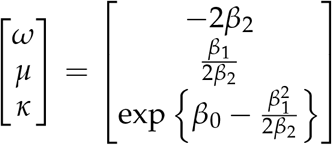

Thus to fit the model, we estimate a hierarchical negative binomial regression model with mean *λ* and overdispersion parameter *r*, as follows:

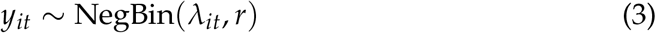

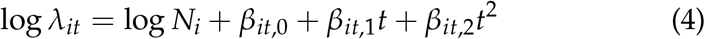

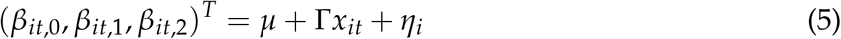

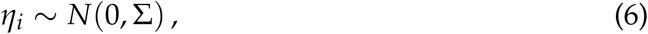

where E(*y*_*it*_) = *λ*_*it*_ and Var(*y*_*it*_) = *λ*_*it*_(1 + *λ*_*it*_/*r*). Here we have included an offset for the logarithm of *N*_*i*_, the population in area *i*, so that the linear predictor can be interpreted as the log per-capita death rate. Here *x*_*it*_ is a vector of social-distancing predictors that are allowed to “flatten the curve” by changing its shape, via the second-stage regression. The negative binomial model naturally handles the heteroscedasticity and overdispersion that we observe in the data. In addition to the polynomial terms in the first-stage regression model for log *λ*, we also include a fixed effect for weekend days, to account for the observed under-reporting bias on weekends.

One note: we have phrased this model as a raw polynomial. But we actually fit the model using orthogonal polynomials to stabilize estimation; one may convert between the raw and orthogonal parameterizations easily.

We fit the model using Markov Chain Monte Carlo, sampling from the posterior distribution of all model parameters. We use weakly informative priors on the fixed effects *β*, the second-stage regression coefficients G, and the covariance matrix S of the random effects. We also explored the possibility of using more informative priors based on daily death rates from European countries. But this had almost no effect on the fit of the model. Model fitting was carried out using the rstanarm package in R [Goodrich et al., 2020]. Forward-looking model projections are based on draws from the posterior predictive distribution. Because of the way that our covariates are constructed (see below), most of the *x* values for these forward-looking projections correspond to social-distancing behavior that has already been observed. However, at a longer horizon some extrapolation of *x* is necessary to generate projections; here we assume that socialdistancing behavior in a state remains unchanged from the average behavior over the seven most recent days of available data.

### 3.3 Social-distancing predictors

To define the social-distancing predictors *x*_*it*_, we take a weighted averages of past social-distancing metrics made available to us by SafeGraph. The metrics considered as predictors include daily time series per location for:

- the median duration of time that people in a given area spend at home, as well as the number of people in an area exhibiting “full-time work” behavior, at their normal place of work;[SafeGraph, 2020b]
- and total *per capita* visitation counts for various points of interest aggregated by category, including grocery stores, hospitals, parks, restaurants, bars, colleges, etc.; [SafeGraph, 2020a]

These data are derived from GPS traces of tens of millions of mobile phones nationwide. “Home” and “work” locations for a device were inferred by SafeGraph based on daytime and overnight locations over an extended period of time. The data were provided to us in an aggregated format; no device-level data or individual GPS traces were accessible by the research team. Figure 2 shows a selection of these social-distancing measures over time in both New York and Texas.

**Figure 2:**
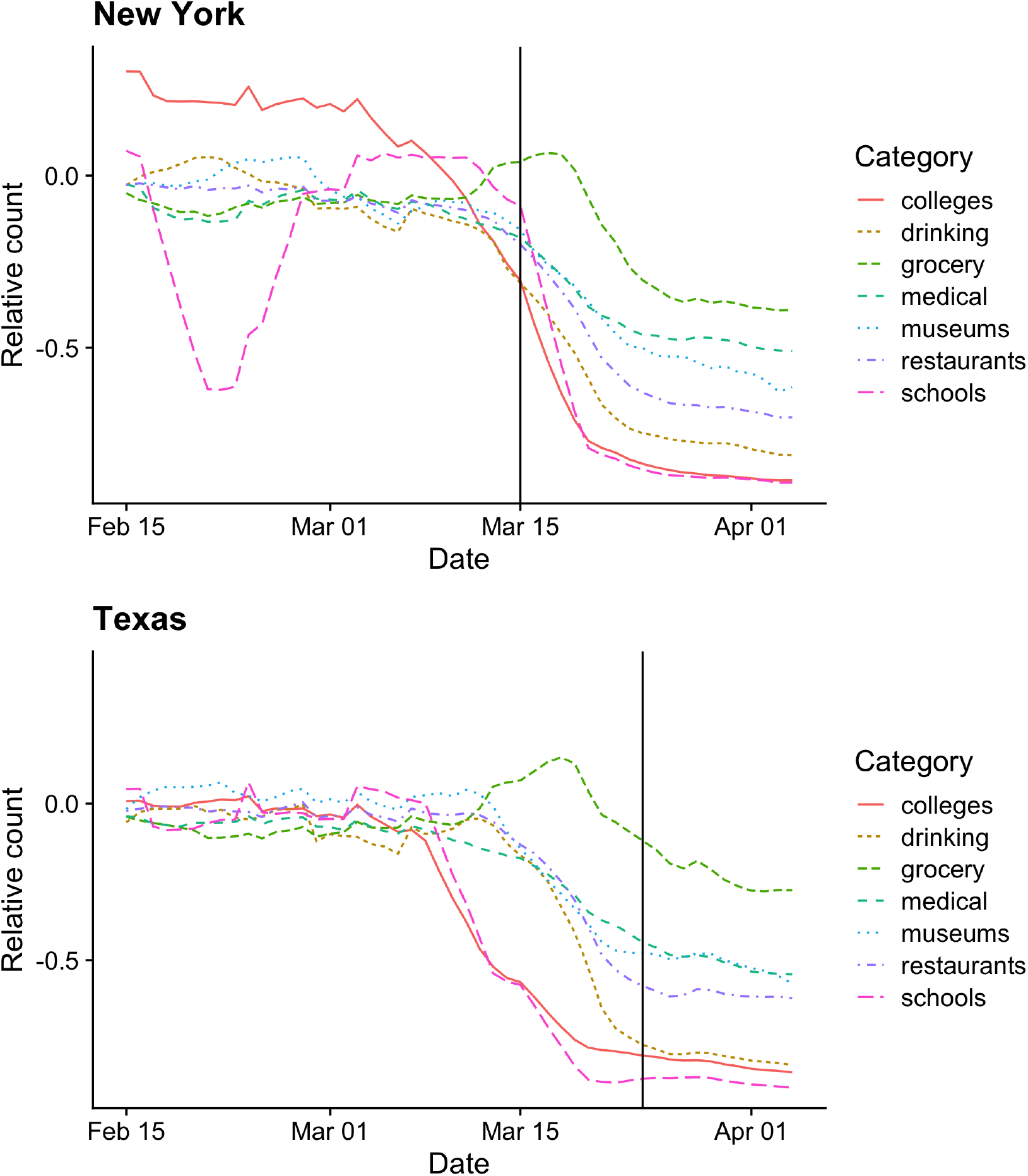
SafeGraph social-distancing data in New York versus Texas. The social-distancing predictors we use in our model quantify visitation patterns to various points of interest, using GPS traces derived from mobile phones. The vertical axis is standardized so that 0 represents a pre-pandemic baseline, and −0.5 indicates a 50% decrease in visitations relative to that baseline. The vertical lines in each panel represent the day in which the state death rate reached 3 per 10 million residents. One can see that social distancing in Texas began substantially before this threshold day was reached, but much nearer this day in New York. Note: the museums category also includes public parks.

We denote these *D* distancing metrics *s*_*it*,1_, …, *s*_*it,D*_, observed each day in each state. To construct useful covariates out of this information, we proceed as follows. For each distancing metric *s*_*j*_, define a corresponding lagged version 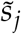 as follows:

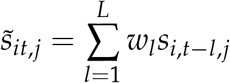

where *w* = (*w*_1_, …, *w*_*L*_) is a fixed vector of backward looking weights. We construct two sets of predictors in this manner. The first uses a weight vector *w*^(1)^ tuned based on prior knowledge of the distribution of lags between infection with COVID-19 and death. Specifically, *w*^(1)^ is a Gaussian kernel centered 23.5 days in the past, with a standard deviation of 6 days. This is based on published estimates of time from contraction to illness onset [Lauer et al., 2020] and on time from illness onset to death [Zhou et al., 2020]. The second uses a weight vector *w*^(2)^ with equal weights over the previous most recent 7 days. Using both sets of weights in our model allows the shape of the curve to adapt not just to the level of social distancing, but to changes over the past month.

To construct predictors *x* for our regression model, we apply principal components analysis (PCA) to two groups of predictors corresponding to the two weight kernels *w*^(1)^ and *w*^(2)^ respectively. For *w*^(1)^ (23.5 days centered Gaussian kernel) we use the first 3 components. For *w*^(2)^ (7-day running average) we use only the first principal component. It is sensible to allocate more predictors from the 23.5 days lagged kernel group since it is motivated by previous research and the known dynamics of contagion. In practice, adding the 7-day running average improves the forecasting performance.

## 4 Results

We update our model results daily and post them at https://covid-19.tacc.utexas.edu/projections/.

To assess forecasting accuracy, we fit our model using data up through April 3rd. We then compared our model’s projections over the next 13 days with those of the IHME model. Figure 3 shows the projections for Texas, California and New York compared with the atual observed deaths in the forecasted period. Our 95% Bayesian credible intervals contain 98.4% of the data points across all states in the heldout data. Similarly, Figure 4 shows the forecasts for the same period for the smaller states. The figure shows that our model remains well calibrated even with few highly dispersed observations. Overall, our model significantly outperforms the IHME model in out-of-sample accuracy, achieving 42% lower 13-day-ahead prediction error: 13.2 deaths per day mean absolute error across all U.S. states, versus 22.8 deaths per day mean absolute error for the IHME model.

**Figure 3:**
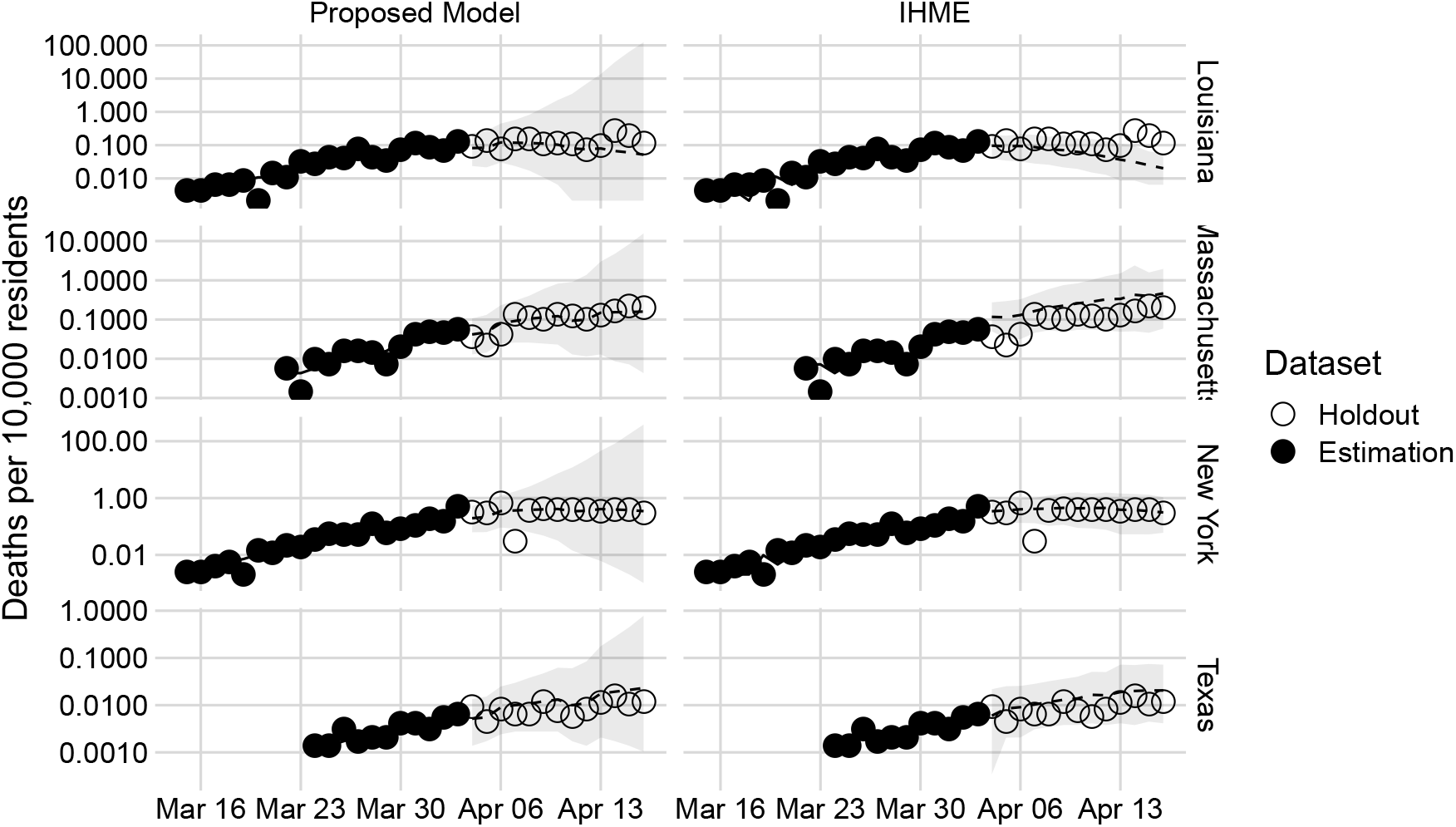
Out-of-sample projections 13 days ahead (log-scale) for Louisiana, Massachusetts, New York, and Texas. The figure shows a comparison of the projections using data up through April 3rd for the 13 days days up through April 16th. We chose 13 days rather than 14 days ahead because the IHME forecasts were publicly available from April 3 but not April 2. Credibility intervals of 95% are shown in gray.

**Figure 4:**
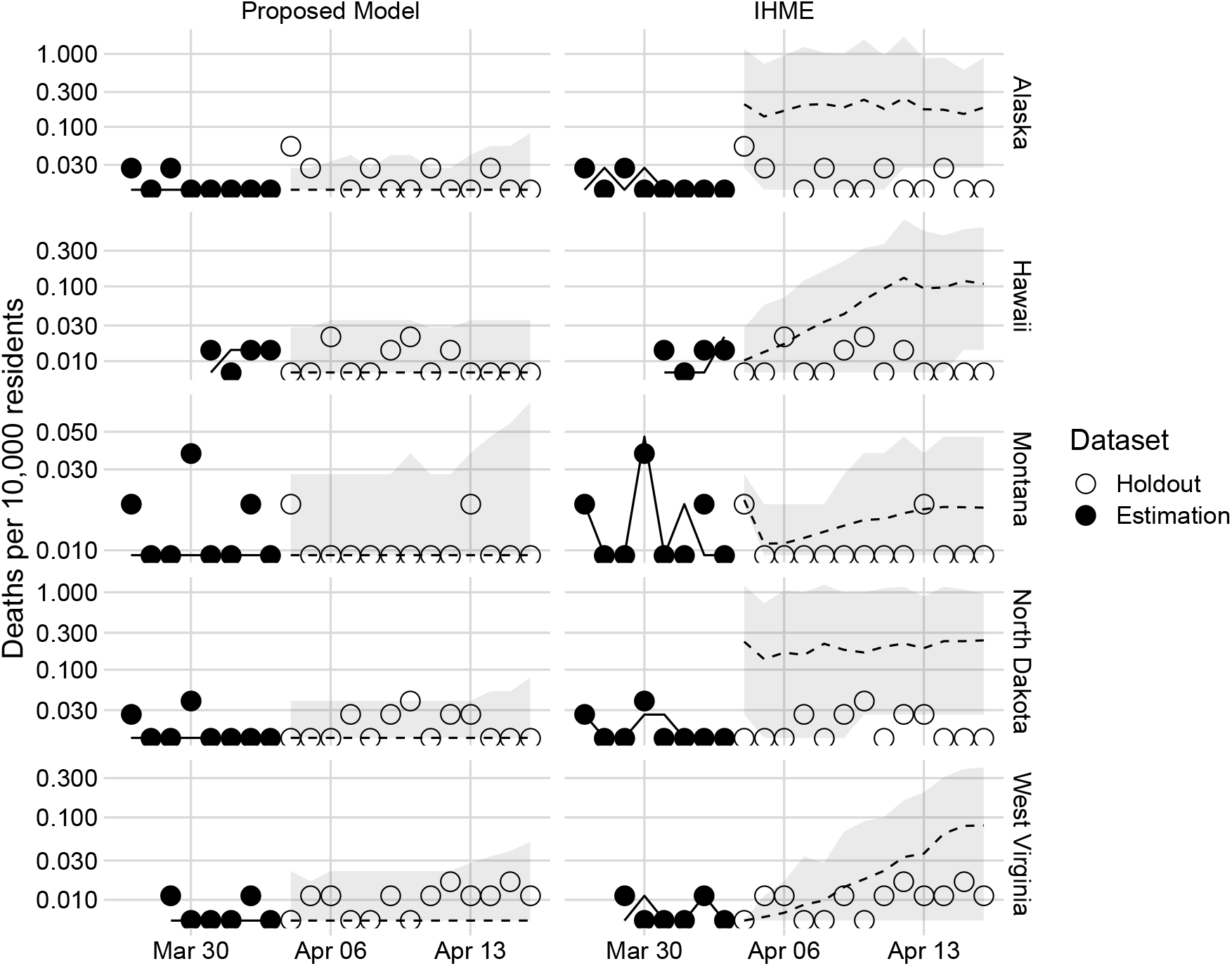
13 day forecast of COVID-19 deaths (log-scale) for a selection of states with fewer overall deaths. The figure shows a comparison of the forecasts using data up to April 3rd for the following days up to April 16th. Credibility intervals of 95% are shown in gray.

### A Uncertainty quantification in the original IHME model

In the IHME model, the choice of using penalized-least-squares fit on the logcumulative-deaths scale has major consequences for statistical inference. In particular, the authors use the inverse-Hessian matrix at the MAP estimate in order to produce uncertainty estimates. This uncertainty quantification procedure, however, implicitly assumes that successive observations are independent. Indeed, without this assumption, it is not generally true that the inverse-Hessian at the MAP provides a valid large-sample estimate for the covariance matrix of an estimator, Bayesian or otherwise. This important technical condition simply cannot be true on the scale used for fitting the IHME model, for the simple reason that the data used for fitting are cumulative: if today’s prediction for cumulative death rate is too high, then tomorrow’s prediction is more likely to be too high as well.

This is easily verified by a simple calculation. The covariance of two successive cumulative death rates *r*_*it*_ is:

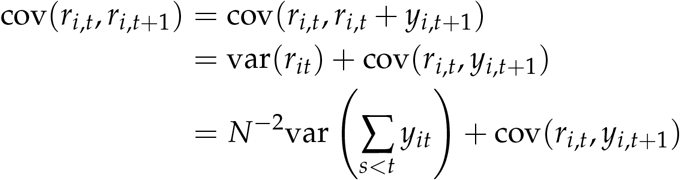

This covariance cannot be zero unless cov(*r*_*i,t*_, *y*_*i,t*+1_) is negative, and of sufficient magnitude to exactly cancel out the first (positive) term—which is highly implausible. Moving to the logarithmic scale does not repair the basic fallacy of assuming independent errors. This likely accounts for much of the understatement in uncertainty seen in Figure 1.

## Data Availability

Data on COVID-19 deaths were provided by the New York Times and are publicly available: https://github.com/nytimes/covid-19-data/ Data on social-distancing metrics were provided under a Data Use Agreement by SafeGraph and are available by contacting SafeGraph directly: https://www.safegraph.com/dashboard/covid19-shelter-in-place

https://www.safegraph.com

